# Undetectable=untransmittable (U=U) messaging increases uptake of HIV testing among men: Results from a pilot cluster randomized trial

**DOI:** 10.1101/2020.11.23.20236695

**Authors:** Philip Smith, Alison Buttenheim, Laura Schmucker, Linda-Gail Bekker, Harsha Thirumurthy, Dvora L. Joseph Davey

## Abstract

HIV testing coverage in sub-Saharan Africa is lower among men than women. We investigated the impact of a peer-delivered U=U (undetectable equals untransmittable) message on men’s HIV testing uptake through a cluster randomized trial with individual mobile clinic days as unit of randomization.

On standard of care (SOC) days, peer promoters’ informed men about the availability of HIV testing at the mobile clinic. On intervention days, peer promoters’ delivered U=U messages. We used logistic regression adjusting for mobile clinic location, clustering by study day, to determine the percentage of invited men who tested for HIV at the mobile clinic.

Peer promoters delivered 1048 invitations over 12 days. In the SOC group, 68 (13%) of 544 men invited tested for HIV (3, 4.4% HIV-positive). In the U=U group, 112 (22%) of 504 men invited tested for HIV (7, 6.3% HIV-positive). Men in the U=U group had greater odds of testing for HIV (adjusted odds ratio=1.59, 95% CI=0.98-2.57).

Tailored, peer-delivered messages that explain the benefits of HIV treatment in reducing HIV transmission can increase men’s HIV testing uptake.

## Introduction

In South Africa and many other countries in sub-Saharan Africa (SSA), men are less likely than women to know their HIV status, start and continue on antiretroviral therapy (ART), and have increased risk of mortality resulting from AIDS-related illnesses (1–5). In March 2020, almost 90% of adult South African males living with HIV knew their status (89% vs. 94% of South African females), 66% of those were on treatment (vs. 77% of females), and 79% of those were virally suppressed (vs. 93% of females), falling short of the 90-90-90 targets (5). Among a number of barriers to testing, some that are frequently cited by men include HIV-related stigma and a fear of an HIV-positive result. Interventions that can effectively address men’s fears of HIV testing are needed to increase testing coverage.

Taking daily antiretroviral therapy (ART) eliminates enough of the virus so that HIV cannot be detected via viral load testing (6). Recent studies have demonstrated that HIV-positive individuals with an undetectable viral load cannot transmit HIV to sexual partners or through giving birth (6,7), a message that is commonly referred to as U=U (undetectable equals untransmittable). As a health promotion message, U=U encapsulates two important aspects of HIV care: knowing your status can bring peace of mind; and starting and staying on ART improves your health and alleviates the worry of HIV transmission to sex partners. The U=U message has been effectively promoted in Europe and North America where people living with HIV describe feeling free of internal and external stigma as a result (8). However, studies outside of South Africa suggest low familiarity with the U=U concept (9,10), and little is known within South Africa about either familiarity with or efficacy of the U=U to message to promote HIV care. Rigorous evaluations of interventions incorporating U=U messaging are needed to understand how best to leverage this important advance in treatment-as-prevention to improve HIV testing, treatment and viral suppression in men, particularly in HIV endemic, low- and middle-income countries.

Behavioural economics (11), a field at the intersection of economics and psychology, suggests that health messages can be ineffective if they emphasise disease and vulnerability (12,13). The behavioural economics model suggests that messages may be more impactful if they are reframed to emphasise desirable, positive outcomes. From this standpoint, U=U messaging has the potential to increase HIV testing uptake by deemphasises the virus and emphasizing the ability to continue living a normal life while maintaining health and protecting of one’s family and sex partners. Behavioural economics-informed interventions can often be delivered in the form of nudges such as small changes in the framing of messages that result in behaviour change without excessive constraints on individuals’ ability to make their own choices (14). Approached that use nudges use behavioural insights to develop interventions that can be easily tested in the field without requiring major changes to the implementation of health services.

We developed a user-designed U=U message for South African men and conducted a pilot randomized trial to investigate the impact of these messages, delivered by interpersonal communication, on men’s uptake of HIV testing. We hypothesized that U=U messages would increase HIV testing uptake among men when compared with standard of care invitations for HIV testing.

## Methods

We investigated the effect of a tailored U=U message delivered by trained male peer promoters on men’s uptake of mobile HIV testing services in the Klipfontein Mitchells Plain (KMP) District in Cape Town. Men in the intervention group received U=U messages whereas men in the standard of care received standard of encouragement seek free HIV testing—both from the same peer promoters.

The study began on March 03, 2020. Due to the COVID-19 outbreak and lockdown, the University of Cape Town closed all non-therapeutic studies on March 18^th^, 2020, after 12 study days had been completed at five sites.

### Setting

Our study was conducted in KMP District, a resource-limited, densely populated, high HIV disease burden area in Cape Town, where the use of health services among men is sub-optimal. The Desmond Tutu HIV Foundation (DTHF) Tutu Tester offers mobile HIV testing clinic days in various locations in the region, including KMP. Two trained male peer promoters stationed near the Tutu Tester mobile clinic sites distributed invitation cards inviting men to visit the mobile clinic for a voluntary HIV test.

### Inclusion criteria and recruitment

Peer promoters were trained to deliver brief messages to men at selected high foot-traffic sites in the KMP community. Males ≥18 years of age within the vicinity of the mobile Tutu Tester HIV testing van and willing to consent to participate in the study were eligible for inclusion.

### Design

We used a cluster randomized trial to determine the effect of the U=U message on HIV testing uptake (primary outcome) and HIV positivity (secondary outcome) in adult men. Individual mobile clinic days served as units of randomization. On each mobile clinic day, men in the vicinity of the mobile clinic were offered invitation cards for same-day testing (**Figure 2**). The information content on the referral cards and the mobilization approach depended on the random allocation day for the intervention or control condition. Invitation cards had unique identification codes. Computer-generated randomization was performed to determine study group assignment for each clinic day, and randomization was stratified by testing location so that an equal number of clinic days at each location within KMP were assigned to each study group.

**Figure 1.**
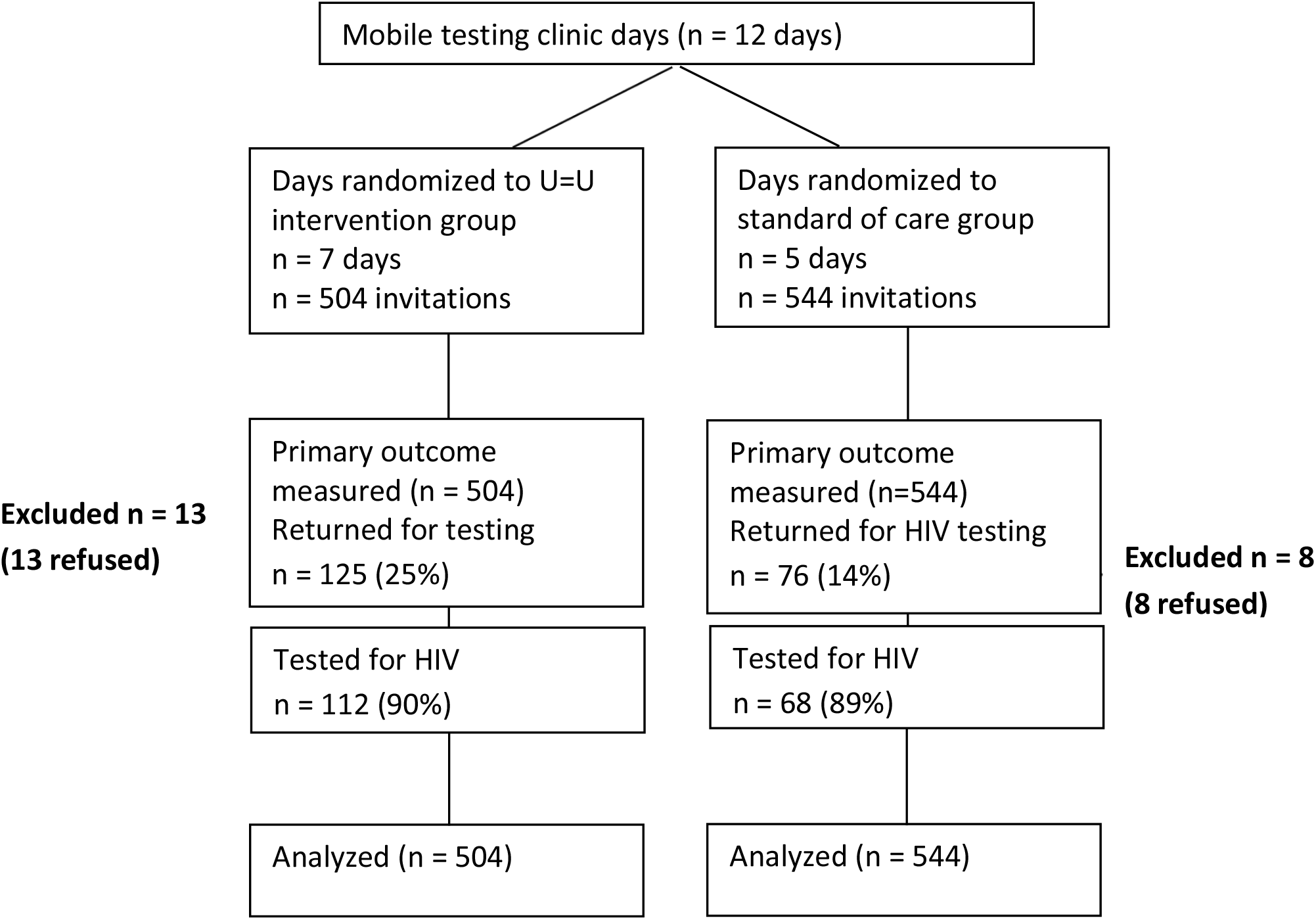
CONSORT Diagram (24)

**Figure 2.**
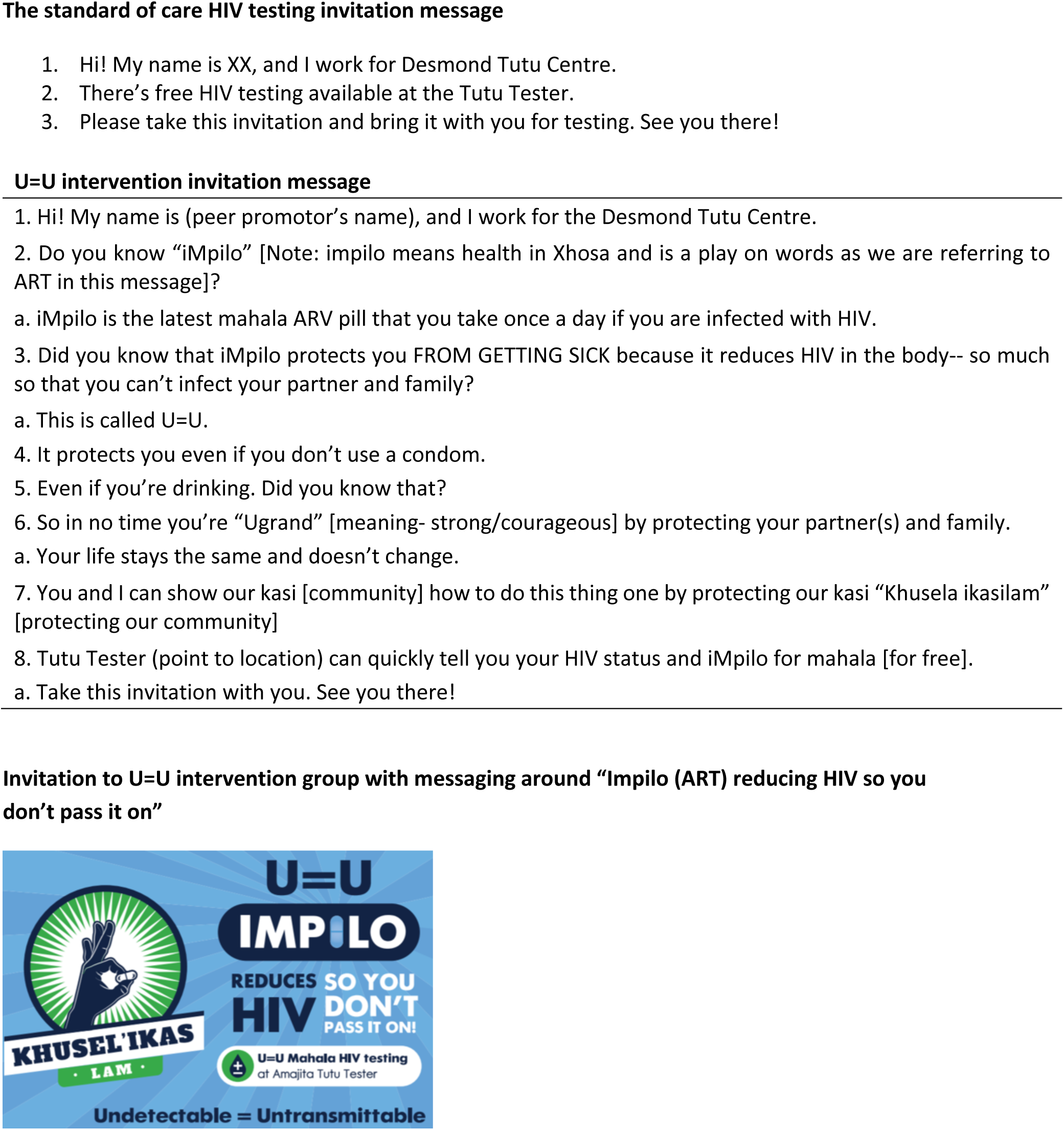
Invitation messages delivered by HIV testing peer promoters.

### Interventions

We used a human-centred design process to develop the U=U messaging (15–19). In collaboration with a local human-centered design company, we conducted two workshops in the local language, isiXhosa, with 50 men from KMP District. Using insights about U=U that were co-discovered with workshop participants, we iteratively developed, piloted, and refined brief messaging about U=U that could be delivered in a face-to-face encounter to encourage same-day testing at the nearby mobile clinic. The final intervention was a 30-40 second script that emphasized eight key insights including the core U=U message **(Figure 2**): 1) the name and workplace of the peer promoter, 2) the colloquial name of ART, as defined by men during the workshop (*Impilo)*, 3) colloquial U=U message, 4-7) male health prioroties (including that your life won’t change if you are living with HIV), 8) directions to the mobile testing unit. Overall, the intervention sought to assuage the fears of testing HIV-positive by conveying the message that HIV treatment makes it possible for people living with HIV (PLHIV) to live a normal life, be healthy, and prevent onward HIV transmission. The U=U (intervention) script was delivered by two trained peer promoters.

The standard of care message promoting HIV testing was delivered by the same two peer promoters and consisted of a 10-second script inviting men to free HIV testing to “know your status” at the nearby Tutu Tester. Each message (intervention and standard of care) was printed on an invitation card that men were told to present at mobile testing sites.

### Process

When men presented their invitation card at the Tutu tester for testing, reception staff outside the testing van collected the invitation and recorded the date and time of presentation. If there were others waiting, men had to wait in line to test. At that point staff reported that some men left because of the waiting time. When it was their turn, a trained HIV counsellor invited the participant into a cubicle in the mobile clinic to complete a rapid HIV test and counselling. The counsellor conducted an individual voluntary written informed consent with the participant and any record reasons for non-participation. Key information collected from participants who consent to participate in the study includes demographic information (age, education, employment), prior testing, relationship status, alcohol use (AUDIT-C) (20–22), prior knowledge of U=U and how U=U affected HIV testing and post-test behaviour including ART uptake. Those testing HIV-positive received post-test counselling and referral to their preferred clinic facility for ART initiation. Participants who were referred for ART were followed up until they started ART.

### Sample size and power

The Tutu Tester regularly visits five sites in the KMP region and serves one site per day. These five sites were randomised to intervention or standard of care days. On a given clinic day, each peer promoter was asked to invite 50 men to test (total of 100 men per day). A sample size of 40 clinic days would result in 4000 invitations (2000 per study group) and result in 80% power to detect a difference in HIV testing uptake of 5 percentage points between intervention and standard of care days, assuming a baseline reporting proportion of 8% testing uptake that was observed during a two-day pilot with the standard of care invitation.

### Analyses

We determined the effect of the U=U intervention on uptake of HIV testing using a logistic regression model that included location fixed effects and standard errors that were adjusted for clustering by study day. We also compared the demographic and behavioural characteristics of the men who accepted HIV testing by study group using chi-squared and t-tests.

### Ethics

The study was reviewed and approved by the IRB at the University of Cape Town (reference HREC ref 750/2019). The study was registered at clinicaltrials.gov under NCT04364165.

## Results

Between March 3, 2020 and March 18, 2020, peer promoters delivered 1048 invitations over 12 days (mean=87 invitations/day). The peer promoters delivered 544 invitations on the standard of care (SOC) days, and 504 on the U=U intervention days, averaging 108 per day in SOC days, and 72 per day in the U=U days (**Figure 1)**.

Demographic and socio-economic characteristics of participants who tested for HIV were similar in the two study groups **(Table 1)**. The average age of those who tested was 36.5 years. Employment characteristics differed significantly among testers in the two study groups, with those in the SOC group having higher employment (66% vs 49%, p=0.03). Most men had tested for HIV before (96%), over half the men were married or cohabiting (57%), and 62% reported recent hazardous alcohol consumption.

**Table 1.** Demographic and HIV risk factors in men testing in Tutu Tester by U=U intervention vs. standard of care arm in Cape Town, South Africa (Feb-Mar, 2020)

|  | **Total**<br>**(n=180)** | **Standard of care**<br>**(n=68)** | **U=U intervention**<br>**(n=112)** | **p-value** |
| --- | --- | --- | --- | --- |
| Age (mean, SD) | 36.5 (12.2) | 36.2 (12.1) | 36.7 (12.4) | 0.31 |
| Education (% completed secondary or above) | 62 (34%) | 23 (34%) | 39 (35%) | 0.89 |
| Employed | 100 (56%) | 45 (66%) | 55 (49%) | **0.03** |
| Monthly income (>\$200/m; R3000) | 75 (42%) | 34 (50%) | 41 (37%) | 0.06 |
| Informal housing | 87 (48%) | 35 (51%) | 52 (46%) | 0.51 |
| Water in home | 113 (63%) | 46 (68%) | 67 (60%) | 0.29 |
| Current relationship status |  |  |  | 0.61 |
| Married/cohabiting | 102 (57%) | 40 (59%) | 62 (55%) |  |
| Single | 64 (36%) | 25 (37%) | 39 (35%) |  |
| Other (divorced, widow) | 13 (7%) | 3 (4%) | 10 (9%) |  |
| Prior HIV test | 173 (96%) | 63 (93%) | 109 (97%) | 0.14 |
| Partner HIV test | 73 (41%) | 29 (43%) | 44 (39%) | 0.66 |
| Number of sex partners in past 6m (mean, SD) | 1.5 (0.96) | 1.5 (0.94) | 1.5 (0.98) | 0.79 |
| Ever exchanged gifts, money for sex | 5 (3%) | 2 (3%) | 3 (3%) | 0.92 |
| Hazardous consumption of alcohol (6+ drinks monthly or more) | 111 (62%) | 40 (59%) | 70 (63%) | 0.58 |

Overall, 125 men returned for testing in the U=U group (25%) vs. 76 (14%) in the SOC group (p<0.01). Twenty-one men left the study van before testing, most likely due to long wait times (n=13 in U=U group and n=8 in the SOC group). In total, 180 (17%) tested for HIV (mean age=37 years). In the SOC group, 68 (13%) of 544 invited were tested for HIV and 3 (4.4%) tested HIV-positive. In the U=U group, 112 (22%) of 504 men invited were tested for HIV and 7 tested HIV-positive (6.2%). Compared with participants in the SOC group, men in the U=U study group had greater odds of coming into the mobile clinic (aOR=1.61, 95% CI=0.99, 2.60) and to get tested for HIV (aOR=1.58, 95% CI=0.98, 2.57) (**Table 2)**.

**Table 2.** Logistic regression models to evaluate the effect of U=U messaging on men returning to test, HIV testing, positivity and linkage to ART.

|  | **Standard of care** | **U=U intervention** | **OR (95% CI)\*** | **aOR (95% CI)\*** |
| --- | --- | --- | --- | --- |
| Invited and came for HIV testing | 76 (14%) | 125 (25%) | 2.03 (1.48, 2.78) | 1.61 (0.99, 2.60) |
| Invited and tested for HIV | 68 (13%) | 112 (22%) | 2.00 (1.44, 2.78) | 1.59 (0.98, 2.57) |
| Tested HIV-positive | 3 (4.4%) | 7 (6.23%) | 1.44 (0.36, 5.78) | 1.60 (0.40, 6.47) |
| Linked to ART | 2 (67%) | 3 (43%) | 0.41 (0.01, 7.81) | -- |

\*Both models included clustering on study day; adjusted model included location

Five (50%) the 10 men who tested HIV-positive were linked into care within six weeks of testing at the mobile clinic (confirmed with National Health Laboratory Service records). In the SOC group, two of three (67%) linked to care. In the U=U group, two of the seven men linked to care, one was already in care before enrolling in the study (43%), and four were not linked to care within six weeks. Positivity and linkage to ART did not differ by group (aOR for positivity=1.60, 95% CI= 0.40, 6.47; OR for linkage= 0.41, 95% CI=0.01, 7.81).

When asked about their beliefs about HIV transmission, most participants who tested agreed or strongly agreed that ART could reduce onward HIV transmission (92%), that a viral load test measured the amount of HIV in one’s blood (88%), and that those with an undetectable viral load could not transmit HIV (80%) **(Table 3)**. Almost three quarters (71%) reported prior knowledge of the U=U message (60% in SOC group, 77% in U=U group; p=0.017). Over two thirds (70%) of participants in the U=U group stated that they heard the U=U message from the peer promoter, followed by the clinic (14%) and family/friends (9%). When asked if the peer promoter had shared the U=U message, almost half (40%) of those in the SOC group and over two thirds (70%) in the U=U group responded affirmatively. Most men in the U=U group (94%) responded that the U=U message encouraged them to test and disclose their HIV status (88%). When asked how they felt after hearing the U=U message, over half stated that they felt relieved (55%) or confident to test for HIV (15%).

**Table 3.** HIV beliefs and encouragement to test in men who tested in mobile tester by study arm, Cape Town, South Africa (Feb-Mar 2020)

|  | Total<br>(n=180) | Standard of<br>care (n=68) | U=U intervention<br>(n=112) | p-value |
| --- | --- | --- | --- | --- |
| Beliefs about HIV transmission |  |  |  |  |
| If partner is HIV+, likelihood of infection is very likely | 140 (78%) | 57 (84%) | 83 (74%) | 0.21 |
| ART can reduce infectiousness of HIV (strongly agree-agree =>5) | 166 (92%) | 64 (94%) | 101 (90%) | 0.36 |
| Viral load measures amount of HIV in blood (strongly agree-agree =>5) | 160 (88%) | 64 (94%) | 95 (85%) | 0.06 |
| Those who have low VL cannot transmit HIV (strongly agree-agree =>5) | 145 (80%) | 53 (78%) | 94 (84%) | 0.57 |
| Heard of U=U before? | 126 (70%) | 41 (60%) | 85 (76%) | **0.02** |
| Where heard it (n=126 who heard of U=U before) |  |  |  |  |
| Peer promoter | 72 (57%) | 11 (27%) | 61 (72%) | **0.01** |
| Family/friend | 18 (14%) | 10 (24%) | 8 (9%) |  |
| Clinic | 27 (21%) | 15 (37%) | 10 (12%) |  |
| TV/Radio | 11 (9%) | 5 (12%) | 4 (5%) |  |
| Other |  |  | 2 (2%) |  |
| Did peer promoter tell you about U=U? (n=126 who heard of U=U before) | 81 (64%) | 16 (39%;<br>13% of total) | 63 (74%, 50%) | **<0.0001** |
| Did information about U=U (n=85): |  |  |  |  |
| Encourage you to test? |  |  | 80 (94%) |  |
| Encourage you to disclose your HIV status? |  |  | 76 (89%) |  |
| How did this information about ARVs reducing HIV in the body so much so that you can't infect your partner make you feel? (n = 112 in intervention) |  |  |  |  |
| Relieved |  |  | 59 (53%) |  |
| Confused |  |  | 2 (2%) |  |
| Confident to test |  |  | 18 (16%) |  |
| Need more information |  |  | 6 (5%) |  |
| No feeling/don't know |  |  | 26 (23%) |  |

## Discussion

Delivery of peer-delivered messages about U=U to adult men in an high HIV prevalence South African setting almost doubled the proportion of men who came to a mobile clinic for free HIV testing. The effectiveness of the brief messages delivered through interpersonal communication in a high-traffic urban area suggests that greater knowledge about the benefits of ART can motivate men to seek HIV testing. Men who were randomized to receive the U=U messages and sought testing had higher levels of knowledge about U=U than men in the SOC group who sought testing, though these men also reported knowledge about ART and viral suppression. Sixty percent of men who were diagnosed with HIV effectively linked into care within 6-weeks (one was already on ART). Even though linkage was measured and found to be higher in the SOC study group, there were few HIV positive diagnoses, limiting our ability to draw inferences.

U=U messaging may work to assuage men’s fear of testing HIV-positive and reassure them that an HIV diagnosis does not necessarily require them to alter their lifestyle. In our U=U message development workshops with men from study communities, men highlighted a fear that testing HIV-positive would limit their ability to have girlfriends, to drink alcohol, and would result in rejection by community members. The U=U message doubled the proportion of men testing in a high HIV prevalence community setting with a higher HIV positivity (6% vs 4%). Even though men in the U=U study group were more likely to return for an HIV test, there was limited difference between the two groups on U=U knowledge. This may suggest a selection effect in which the U=U message motivates testing *without* the participants having to cognitively engage with or recall the specifics of the messages. Moreover, the difference in employment between the two groups (higher in the SOC group) may in some part be due to promoters not having sufficient time to complete the delivery of the U=U invites with employed men because they may have less time than unemployed men to attend to the message and get tested.

Early studies on the U=U message in SSA have found varying levels of awareness or understanding of the extent to which ART reduces HIV transmission risk. Some studies that measured men’s beliefs about the reduction in HIV transmission risk with ART suggest they may underestimate the benefits of ART (23), while others show awareness that consistent ART use stops onward transmission. Overall comprehension of the U=U message and consequent implementation in SSA remains low, however, and there have been limited efforts to translate these messages widely in high HIV burden settings in SSA (10). This study’s findings point to the positive influence that user-designed U=U messages that are adapted to local context may have on men’s decisions to test and link to care. Further work is required, however, to better understand message comprehension and beliefs about U=U. Although our findings indicate that the tailored nudge increased testing uptake, it may be instructive to identify factors, including cognitive factors, associated with message comprehension and subsequent progression through the treatment cascade.

There were a few limitations in this study. The study was cut short due to the 2020 COVID-19 lockdown in South Africa, which ended the study after only 12 days and 1049 participants of the projected 4000. While the study had sufficient power to assess the primary outcome, we may have had more data to evaluate the understanding of U=U and viral suppression on HIV transmission. Further, there may be alternative and potentially more efficient delivery methods in the field including mixed media to include social media, clinic based promotions or SMS messages to build trust around U=U in the community. While face-to-face delivery increased HIV testing in this study, it may be worth investigating other methods to improve men’s HIV testing uptake in South Africa. Future research and implementation efforts should explore most effective, cost-effective ways of delivering accessible information about U=U to men in South Africa and other high HIV prevalence regions in SSA.

This study demonstrated that tailored, peer delivered U=U messaging was an effective nudge which improved testing HIV uptake in adult men, and men with higher HIV positivity. Future studies should investigate how best to scale up delivery of information about U=U in men in the community and clinics to improve HIV testing, treatment and viral suppression. Alternative methods of message delivery, especially considering the limitations on interpersonal interactions during lockdowns, should be investigated and tested. A larger study could assess participants’ understanding of the U=U message, either immediately after hearing the message, or upon HIV testing uptake. Lastly, further research is needed on the impact of U=U on ART uptake, retention and viral suppression in men. Since U=U messaging improved testing uptake, pragmatic implementation methods may hold promise for supporting men through the treatment cascade to achieve viral suppression and require exploration.

## Data Availability

The data are available upon request from the authors.

